# An Epidemic Model SIPHERD and its application for prediction of the spread of COVID-19 infection in India

**DOI:** 10.1101/2020.06.25.20139782

**Authors:** Ashutosh Mahajan, Ravi Solanki, Namitha Sivadas

## Abstract

After originating from Wuhan, China, in late 2019, with a gradual spread in the last few months, COVID-19 has become a pandemic crossing 9 million confirmed positive cases and 450 thousand deaths. India is not only an overpopulated country but has a high population density as well, and at present, a high-risk nation where COVID-19 infection can go out of control. In this paper, we employ a compartmental epidemic model SIPHERD for COVID-19 and predict the total number of confirmed, active and death cases, and daily new cases. We analyze the impact of lockdown and the number of tests conducted per day on the prediction and bring out the scenarios in which the infection can be controlled faster. Our findings indicate that increasing the tests per day at a rapid pace (10k per day increase), stringent measures on social-distancing for the coming months and strict lockdown in the month of July all have a significant impact on the disease spread.

## I. Introduction

The outbreak of novel Corona Virus Disease (COVID-19) caused by severe acute respiratory syndrome coronavirus 2 (SARS-CoV-2), originated from a wet market in Wuhan, China, is now widespread in the world and has severely affected many counties including India. The first case of COVID-19 in India was reported on January, 30 2020 at Thrissur, Kerala, in a student who had returned from China. India has reached the fourth position in the world in the number of confirmed COVID-19 cases and presently has 449,613 confirmed cases and 14,162 deaths as of June 23, 2020, which is really an alarming situation. Social distancing is the best method for mitigating this pandemic until an effective medicine or vaccine is invented [1]. The first nationwide lockdown is ordered by the Prime Minister on March 24 for 21 days and further extended the lockdown till May 3 by relaxing certain substantial fields [2]. Later, though the lockdown period is extended to June 30, the freedom is given to the states to impose restrictions assessing the situations in the respective states.

Mathematical modeling and simulation are helpful for predicting the transmission of the epidemic and to implement necessary actions for its control. Mathematical models for the epidemic have a major role to make predictions of the transmission dynamics of the disease and thus assist the authority to take necessary movements for the containment.

Several epidemic models are already reported in the literature, however, the COVID-19 is different type of infection showing certain special characteristics. The transmission of the disease from the persons who are infected without showing any symptoms (asymptomatic cases) is one of the special characteristics of this disease [3] and to be considered in the modeling. Also, the disease can be spread from the infected who is in the incubation period [4]. In this study, we present a new mathematical model named SIPHERD, incorporating the aforementioned characteristics of the COVID-19.

Many mathematical models for the infectious disease spread are reported in the literature, and the classical and widely used method is SIR model described in [5]. An approximate spatial epidemiological model of the COVID-19, is initially proposed in [6] in which the spread of the disease within between and the countries is analysed. The infected and undetected cases and the spread of COVID-19 from those persons are incorporated in [7] but this study is associated with only China.

A modified compartmental SIR model is discussed in [8]. In this study, the total population of the country is divided into eight compartments, and this work is carried out only for Italy, and also, the model does not undertake the purely Asymptomatic cases of infected. A different compartmental model SEIR [9] predicts the dynamics of the transmission of the COVID-19 for certain countries, and the impact of quarantine of the infected persons are also studied in it. Another improved SIR model is depicted in [10], and the time dependency of the parameters of the SIR model is also examined in this. Prediction of transmission of COVID-19 using curve fitting algorithms are reported in [11], [12] and [13].

A stochastic mathematical model is proposed in [14] to analyse the impact of COVID-19 on the healthcare system in India. Assessment of the preventive measures of COVID -19 such as lockdown and prediction of its spread in India is studied in [15]. The SEIR epidemic and regression model is extended for predicting and evaluating the transmission of COVID-19 in India [16]. Progression of the epidemic in India is also determined using the mathematical modelling in [17] and [18].

In India, complete lockdown is limited to the containment zones and hotspots from June 1 on-wards. The forbidden activities in the places outside the containment zones are re-opened in a phased manner with the conditions to follow the standard operating guidelines given by the Health Ministry of India. Inter-state and intra-state travel is allowed in the present situation without any pass/permission, religious places, restaurants, shopping malls and hospitality facilities are allowed to open whereas the educational institutions, entertainment zones, international air travel and railway services remain prohibited as of June 23. Despite all these restrictions, the infection is growing exponentially, and policymakers need to consider different ways for the containment of the disease. The most hit cities in India by COVID-19 pandemic are Mumbai, Delhi and Chennai with a collective population of around 40 million.

In this paper, we bring out different possible ways for better control of the infection spread. We employ an improved mathematical model SIPHERD [19] for the COVID-19 pandemic embedding the purely asymptomatic infected cases and the transmission of the disease from them. The model simulations bring out the efficacy of different ways for the containment, by predicting the total number of active and confirmed cases, total deaths, and daily new positive cases considering various social distancing/lockdown conditions and the number of tests done per day.

## II. Methods

We model the evolution of the COVID-19 disease by dividing the population into different categories as listed below which is described in detail in [19] [20]. The SIPHERD model equations are for the defined entities (S,I,P,H,E,R,D) are a set of coupled ordinary differential equations (1 to 7). The population is divided into different categories, as Susceptible (S), Exposed (E), Symptomatic (I), Purely Asymptomatic (P), Hospitalized or Quarantined (H), Recovered (R) and Deceased (D).

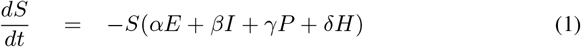

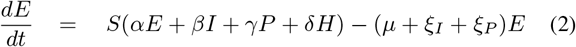

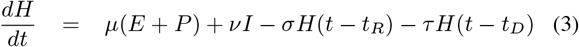

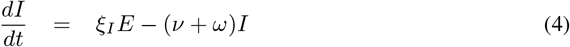

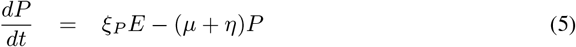

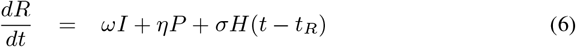

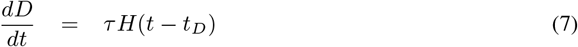

where, *t*_*R*_ and *t*_*D*_ are the delay associated with the recovery and death respectively with respect to active cases *H*. The various parameters seen in Fig 1 and their optimized values for India COVID-19 data are listed in table I.All fractions add up to unity that can also be seen from summing the above equations.

**Fig. 1:**
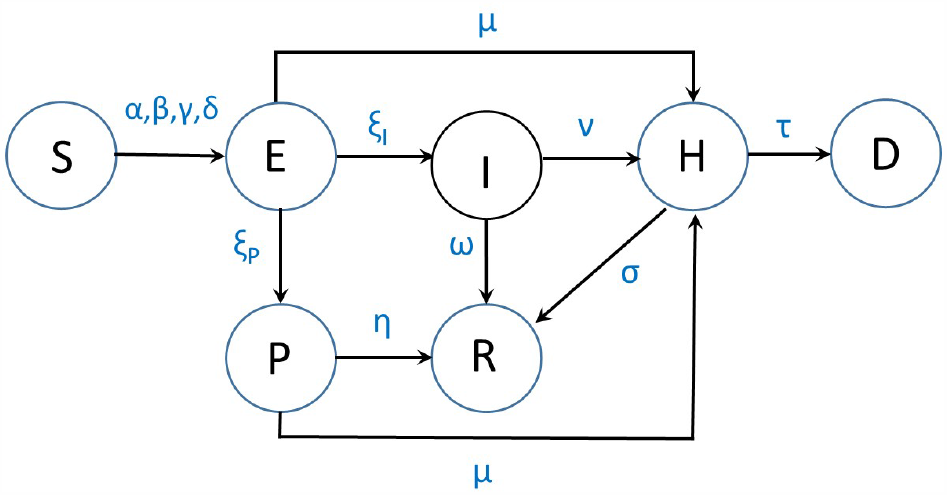
The SIPHERD Model.

**TABLE 1:**
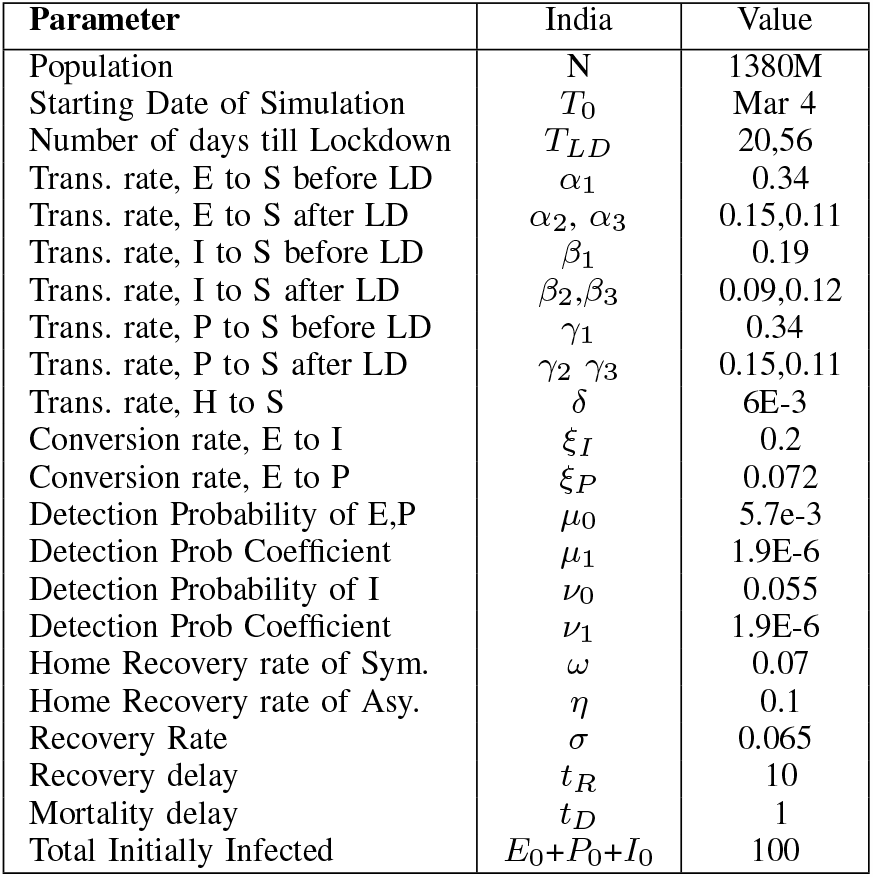
Parameters values for India

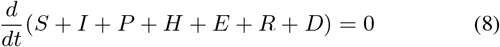

The detection of the Asymptomatic and Symptomatic cases can be taken dependent on the number of tests done per day (*T*_*PD*_).

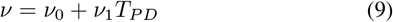

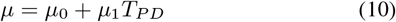

where, *µ*_0_, *µ* _1_, *ν*_0_, and *ν*_1_ are positive constants. The effectiveness of the tests increases if contact tracing is performed. So far in India contact tracing is performed well, we assume that the increased tests are also performed on the suspects more carefully and the detection probability increases with increasing tests linearly. Since the severe cases are going to approach for the tests, one component of the detection probability is not taken dependent on the number of tests.

Recovery of Asymptomatic cases is taken faster than the Symptomatic cases. The total confirmed cases are the addition of the active cases, extinct cases, and a part of the recovered that were detected.

This can be written as

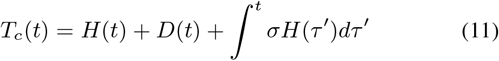

## III. Numerical Implementation and Simulation

The set of coupled ordinary differential equations for the model can be solved numerically for a given set of parameters and initial values. It is however important that the parameters are determined accurately so that the model demonstrates the real situation of the infection spread. We take into account the data sets of the total number of confirmed cases, active cases, cumulative deaths and tests done per day, and find the model parameters that generates the best possible match between the actual data and model. For this purpose, a cost function is written in terms of errors between the actual and solver data sets. The minimizer of the cost gives the optimized set of parameters. The model and the optimization codes are implemented in MATLAB.

## IV. Results and Discussion

The number of total positive or confirmed cases, present active cases and deaths are collected from [21], [22], and the number of tests per day from [23], which is plotted in Supplementary Material Fig.S1 a. The parameters determined by our model for COVID-19 spread in India are listed in TABLE I and the simulation data from the model is compared with the actual data in Supplementary Material Fig.S6. The parameter values related to the characteristics of the disease are discussed in more detail in [19].

### A. Predictions for India

For the available data till June 21, 2020, we run the model for first 109 days i.e. till June 20 to extract parameters listed in TABLE I, and then with the extracted parameters, the model is run for 500 days starting from March 4. The current increase in tests per day is around 2.5k, we assume the same trend for the test per day and take the current value of transmission rate to generate the simulated data for the prediction.

### B. Effect of Lockdown

Two scenarios are considered for lockdown and social distancing conditions. One possible scenario is that the conditions are kept the same, and the second one is that they are made stricter by taking into account some measures after June 21 such that the transmission rates *α* and *β* decrease by 5%. These measures can include restrictions on travel, large gathering of people for social events, distribution of low-cost masks and hand sanitizers in hot spots.

Test per day assumed to be increased by 2.5k, which is close to the current trend and taken saturated at 1 million for both the scenarios. The mortality rate is calculated from the data and is improved in steps from initial value 3.17E-3, 3.14E-3, 2.29E-3, 2.79E-3 on March 4 to June 21 as seen in Supplementary Material Fig.S1.b. For the future, mortality rate is taken improved to a fixed value 2e-3.

A comparison of the predictions for the two scenarios i.e. with and without the stringer measures is plotted in Fig. 2. Reproduction number variation with time and evolution of the undetected infected cases can be seen in Supplementary Material Fig.S2. The total number of reported cases is predicted to be around 12 million. This number appears very high. However, compared to the USA reported cases 2.4 million, the number is reasonable, given the fact that India’s population is roughly four times higher.

**Fig. 2:**
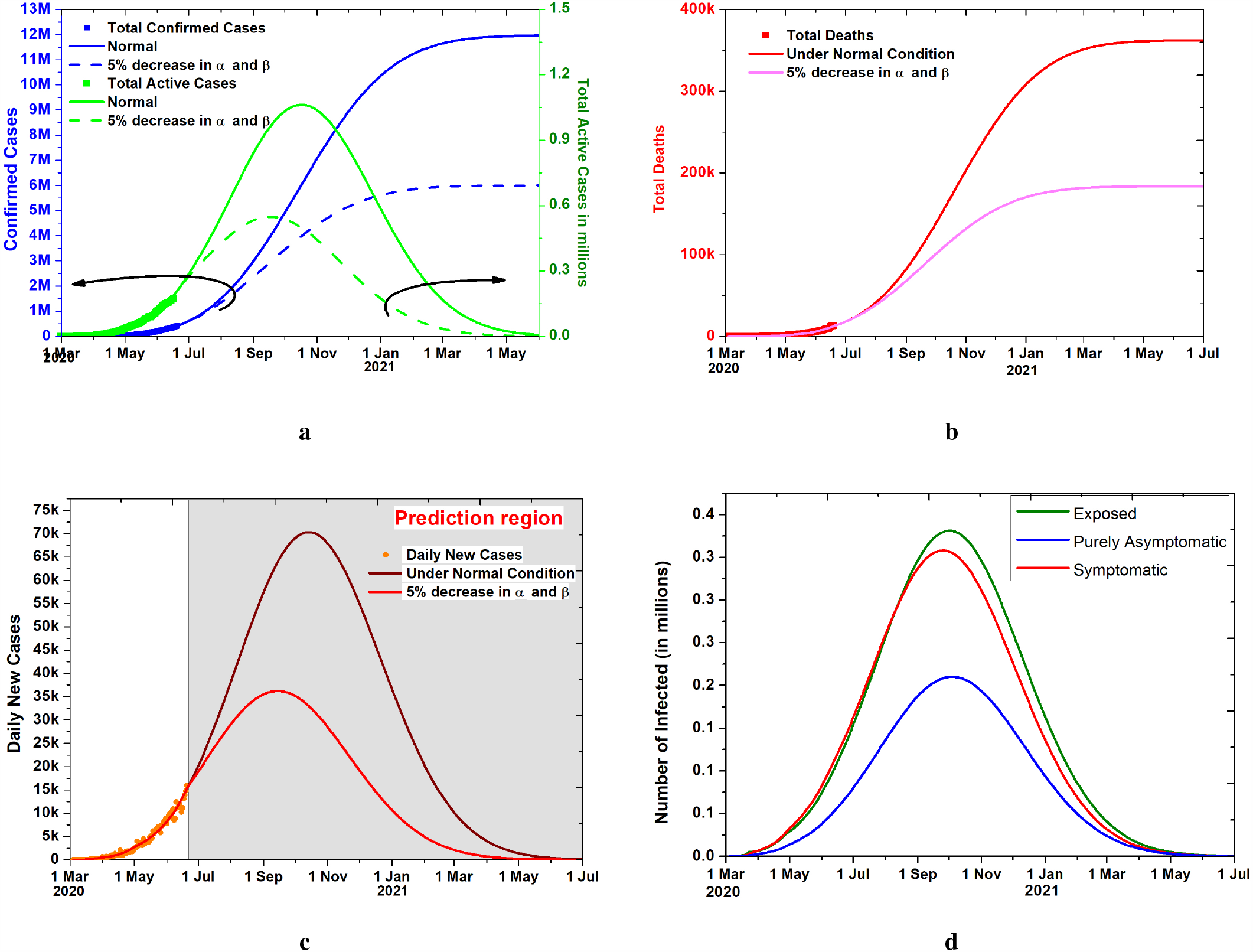
Model predictions for Stringent Measures on Social Distancing. The present social distancing conditions are taken improved further with additional measures such that transmission rates comes down by 5%. **a**,**b**. Model prediction for the confirmed, active and total deaths with and without additional control measures. **c** Comparison of daily new cases in “stringent measures conditions” with the current trend. **d** The evolution of the undetected Exposed, Symptomatic, and purely Asymptomatic cases for 2.5k increase in TPD with social distancing conditions kept the same.

We also study one more possible scenario in which a total lockdown is imposed for July 2020. The rate of transmission of infection is going to decrease in the imposed lockdown, and we take the *α* and *β* values to decrease by 10% from the current value. The prediction with 2.5k increase in tests per day and saturation at 1 million tests, is compared in FIG. 5a,b and in FIG. 5d, we plot the prediction for the daily new cases. Detailed plots and evolution of infected can be seen in Supplementary Material Fig.S5 for this condition of stricter lockdown for July and relaxed after that.

### C. Effect of Increase in Testing

We compare the effect of testing on the prediction of total, active and death cases in FIG. 3 a,b. Total, Active and extinct cases are plotted for the coming months if tests per day are increased by 2.5k, 5k and 10k per day after June 21 and saturated at 1 Million as seen in Fig 4.The factor by which an increase in testing can contain the infection can be estimated.The daily new positive cases data and the prediction for the 2.5k and 10k increase in tests per day are plotted in Fig. 3d.

**Fig. 3:**
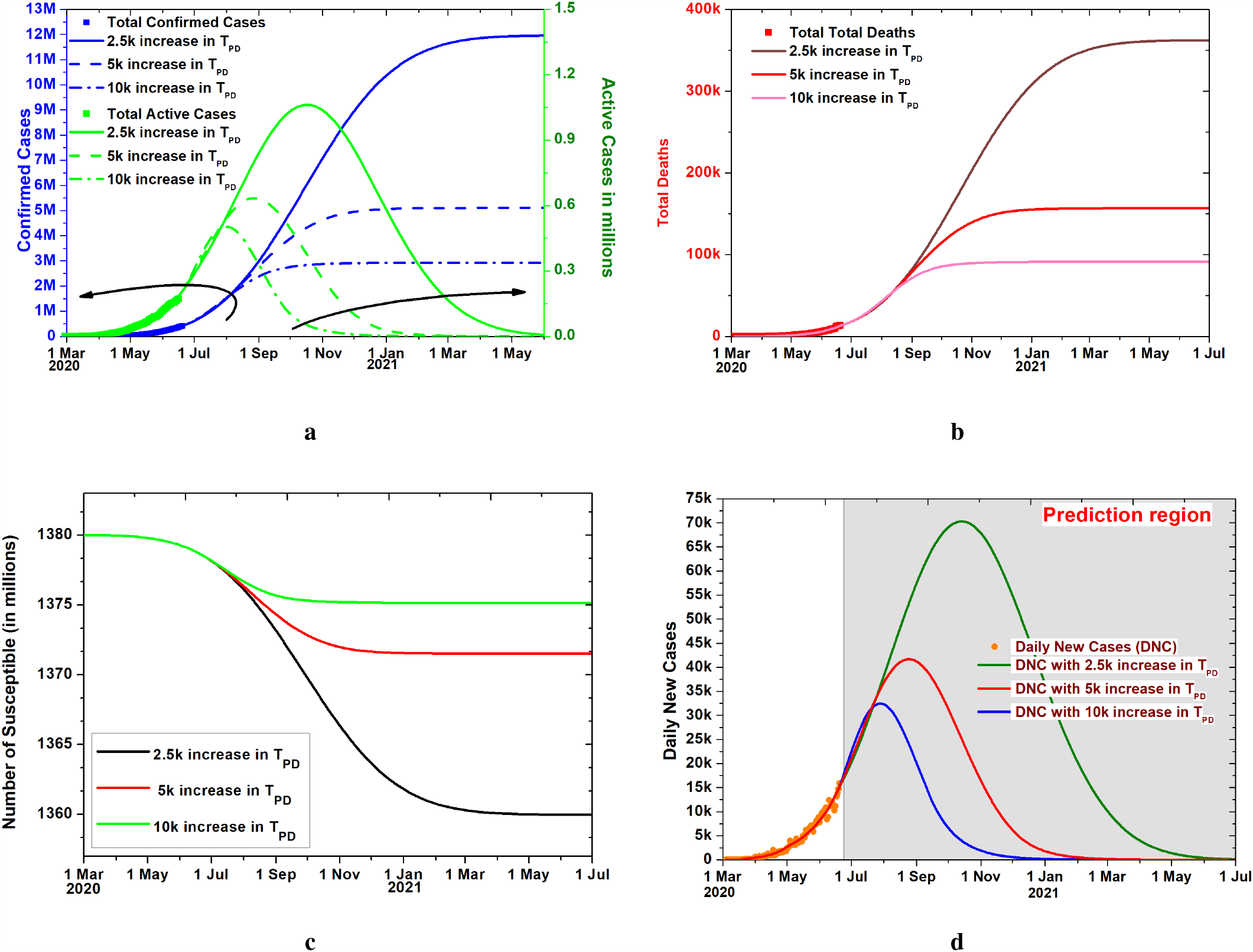
Effect of Increase in Tests. **a**,**b**. Model prediction for the confirmed, active and total deaths cases for different testing conditions. **c**. Evolution of the Susceptible. **d**. The daily new cases for 2.5k, 5k and 10k increase per day in TPD.

**Fig. 4:**
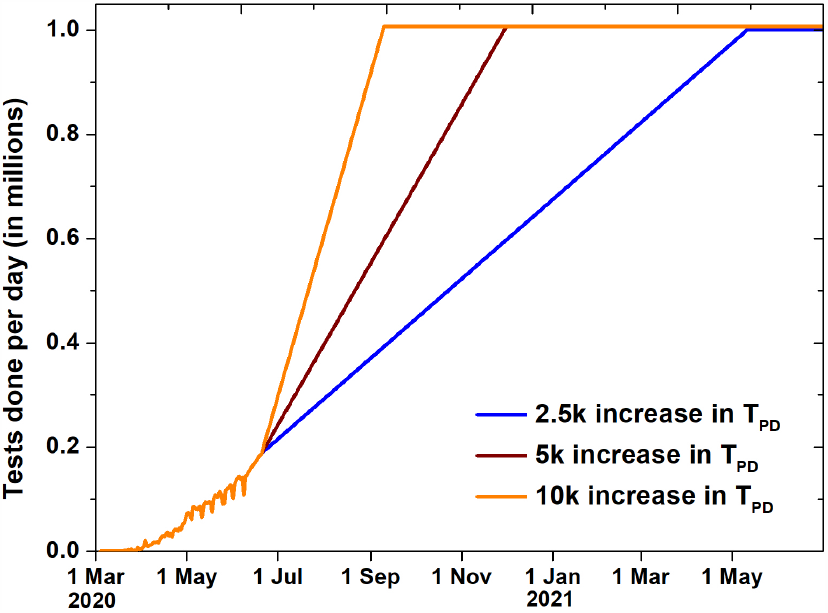
Comparison of 2.5k, 5k and 10k increase in Tests per day (*T*_*P D*_) in future saturated at 1 million.

The initial reproduction number 3.2 is seen go down to 1.5 after the imposition of lockdown on 20th day i.e. March 24. After the second lockdown on 56th day i.e. April 27 it has reached 1.32. It shows a downward trend further and goes below 1 on October 5,2020. If tests per day are increased 10k per day, then the reproduction number comes below one on July, 16, which can also be seen in Fig. 5c.

**Fig. 5:**
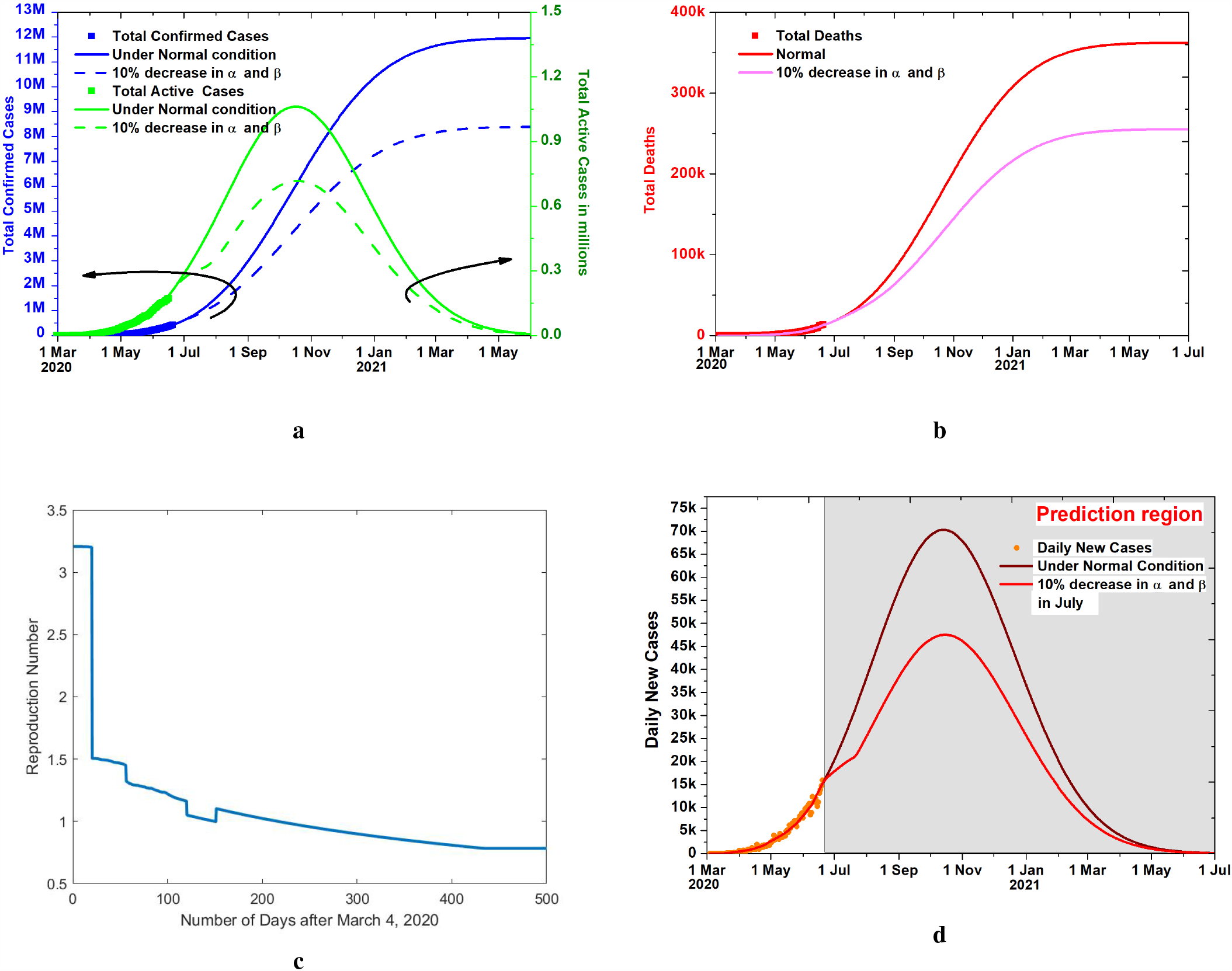
Effect of lockdown in the month of July. **a**,**b**. Effect of one month Lockdown on the total, active and extinct cases, with 2.5k increase in Tests per day (*T*_*P D*_) saturated at 1 million.**c**. Reproduction number variation with number of days. Effect of lockdown can be seen as a dip in reproduction number. **d**. The daily new cases with and without lockdown in July.

The reproduction number is seen to go below one on July 30 if the transmission rate decreases by 10% due to lockdown in July. The initial basic reproduction number 3.2 is in the range of the mean reported value [24] [25].

A sensitivity study is carried out for the different parameters, as seen in Supplementary Material Fig.S4 and S5. The parameters are increased and decreased by 5% from the optimized values to see the changes in the outcomes. It can be seen in Supplementary Material Fig.S5 that the model prediction are most sensitive to the transmission rates *α*_3_,*β*_3_ and *γ*_3_. Just a 5% change in these estimated parameters gives a huge change in total, active and extinct cases. Supplementary Material Fig.S4 show that *ν*_0_ is a sensitive parameter. It indicates that if infected people spontaneously approach the test and quarantine themselves, it will give better control. The fear of testing for the subsequently isolated quarantine if tested positive should be alleviated.

## V. Conclusion

SIPHERD model is employed for COVID-19 spread that considers purely Asymptomatic category of infected cases in addition to the Symptomatic, and the disease spread by the Exposed. The effect of lockdown on the rates of transmission of infection and the influence of tests per day on detection rates has been incorporated in the model. With the current trend, total infections would be 12 million when disease ends and can lead to 362k total deaths. Our findings suggest that increasing the number of tests at 10k per day with highly efficient contact tracing, rather than the current 2.5k per day rise leads to a reduction of 9 million reported cases and reduction of 270k in total extinct cases. In the absence of a vaccine, the infection can last long till the end of this year and number of deaths could be around 350k if social distancing conditions and increase in tests remain at the current trend.

## Data Availability

The number of total positive or confirmed cases, present active cases, and deaths are collected from publicly available data [1], [2], and the number of tests per day from [3].
[1] https://www.worldometers.info/coronavirus.
[2] https://www.covid19india.org.
[3] https://ourworldindata.org/grapher/full-list-covid-19-tests-per-day.

https://www.covid19india.org

https://ourworldindata.org/grapher/full-list-covid-19-tests-per-day

